# Use of amplicon-based sequencing for testing fetal identity and monogenic traits with single circulating trophoblast (SCT) prenatal diagnosis

**DOI:** 10.1101/2020.06.01.20108100

**Authors:** Xinming Zhuo, Qun Wang, Vossaert Liesbeth, Roseen Salman, Adriel Kim, Ignatia Van den Veyver, Amy Breman, Arthur Beaudet

**Affiliations:** Department of Molecular and Human Genetics, Baylor College of Medicine, Houston, TX, USA; Department of Medical and Molecular Genetics, Indiana University School of Medicine, Indianapolis, IN, USA; Graduate Program in Diagnostic Genetics, MD Anderson Cancer Center, Houston, TX USA

## Abstract

A major challenge for cell-based non-invasive prenatal testing (NIPT) is to distinguish individual presumptive fetal cells from maternal cells in female pregnancies. We have sought a rapid, robust, versatile, and low-cost next-generation sequencing method to facilitate this process. Toward this goal, single isolated cells underwent whole genome amplification prior to genotyping. Multiple highly polymorphic genomic regions (including HLA-A and HLA-B) with 10-20 very informative single nucleotide polymorphisms (SNPs) within a 200 bp interval were amplified with a modified method based on other publications. To enhance the power of cell identification, approximately 40 Human Identification SNP (Applied Biosystems) test amplicons were also utilized. This method allowed reliable differentiation of fetal and maternal cells. In fully informative cases, two haplotypes were found within the maternal reads, and fetal cells showed reads with one but not the second maternal haplotype while also showing a novel paternal haplotype absent in the mother. For SNP typing, at least 2 SNPs and 10% of informative SNPs were required to differentiate a fetal cell from a maternal cell. A paternal DNA sample is not required using this method. The assay also successfully detected point mutations causing Tay Sachs disease, cystic fibrosis, and hemoglobinopathies in single lymphoblastoid cells, and monogenic disease-causing mutations in three cell-based NIPT cases. This method could be applicable for any monogenic diagnosis.

## Introduction

Since 2015, several influential professional societies, including the International Society for Prenatal Diagnosis (ISPD) and American College of Obstetricians and Gynecologists (ACOG), have stated that noninvasive prenatal testing (NIPT) is an available screening option for all pregnant women. Current NIPT is based on analysis cell-free fetal (cff) DNA, and it has become widely available since its introduction to clinical practice in 2011. In contrast, cell-based NIPT, which relies on the isolation of circulating fetal cells in maternal blood, has been a long-sought alternative to cell-free NIPT and is now approaching commercialization. Currently, the cell-free NIPT approach has the advantage of a faster turnaround time and lower cost. However, the accuracy of cell-free NIPT is impacted by the large amount of maternal DNA in plasma (more than 80% of all circulating DNA) and the highly fragmented nature of this genetic material. Thus, it is only recommended for detection of the common fetal aneuploidies by many professional societies [1, 2]. These drawbacks can be addressed by cell-based NIPT, but it is not yet available as a clinical test. Cell-based NIPT would potentially have a higher positive predictive value compared to cell-free NIPT, since the DNA source is purely fetal or placental in origin without any maternal contamination. [3-5]. Recently, multiple groups have reported successful cases of cell-based NIPT via capturing trophoblast cells [3, 6, 7].

The critical step for cell-based NIPT is the recovery of rare fetal cells, such as trophoblasts. As described previously [3, 4], 30-40 mL of blood is collected at 10-16 weeks’ gestation, followed by density fractionation or magnetic activated cell sorting (MACS) with anti-trophoblast antibodies to enrich the nucleated cells. Then, the nucleated cells are immunostained to identify trophoblasts that are cytokeratin positive and leukocyte common antigen (CD45) negative. The stained cells are picked individually under fluorescence microscopy with an automatic instrument described previously [3, 4] and subjected to whole genome amplification (WGA), which allows downstream genotyping, and copy number analysis using array Comparative Genomic Hybridization (CGH) or next generation sequencing (NGS).

Genotyping is an essential step after isolating the putative fetal cells. Typically, a successful cell-based NIPT would isolate 5-10 cells per 30 mL maternal blood sample. Since the nucleated cell recovery is a complicated multiple-step procedure, and several antibodies are used, there is a chance of picking a maternal cell (~10% in our experience). Whole genome shotgun (WGS) sequencing at low coverage (5-10 million reads per cell) provides good copy number data, but it does not readily distinguish fetal and maternal cells if the fetus is female. Previously, we used short tandem repeat analysis, SNP arrays, or Y-chromosome targeted qPCR to confirm the fetal origin of single cells. However, there are various disadvantages to these approaches, such as inefficiency, ambiguity, high cost, or limited application.

In this work, we developed a fast, low-cost, and reliable genotyping assay with amplicon sequencing. We sequenced approximately 90 highly polymorphic SNPs within about 40 amplicons. Among these amplicons, four contain multiple common SNPs (see Materials and Methods), which allow for haplotyping of the WGA DNA product. Together, it allows for the effective differentiation of cells containing the fetal genome from cells of maternal origin in most cases. This method could also be easily expanded for the detection of additional disease-associated variants, which would have clinical utility for pregnancies with increased risk for monogenic disorders.

## Materials and methods

### Sample collection and preparation

Blood samples were collected from pregnant women from multiple centers under a protocol approved by the Baylor College of Medicine Institutional Review Boards. Approximately 30 mL of blood was collected into anticoagulant EDTA Vacutainer tubes (BD). Fetal cells were enriched with methods described in Breman et al., 2016. Both cytokeratin (CK)-positive putative fetal cells, and CK-negative maternal white blood cells were picked from maternal blood using the CytePicker® equipment (Rarecyte).

### Overall strategy

The typical cell-based NIPT workflow yields 3-10 singlet or doublet cells per patient, individually captured in a PCR tube for downstream WGA. The WGA DNA products of those cells must be checked for quality and confirmed as nonmaternal cells before finalizing the interpretation. Thus, a fast, low-cost, and high-throughput genotyping assay is necessary. To meet this need, we designed a single-cell genotyping assay using a modified amplicon sequencing approach for genotyping (Fig 1). The first step is conventional PCR with a pool of amplicons with bridging adaptors, which contains a partial sequence of Illumina i5 and i7 adaptors. The second step is adding a dual index with a sequencing adaptor to previous PCR products. This concludes the library construction for the Illumina machine. DNA samples with different indexes were balanced and pooled for sequencing with Illumina Miseq. The sequencing result was demultiplexed with Illumina BaseSpace. The demultiplexed reads were mapped with conventional BWA-MEM. The mapped reads were used for SNP typing and amplicon haplotyping, the details of which are discussed below.

**Fig 1.**
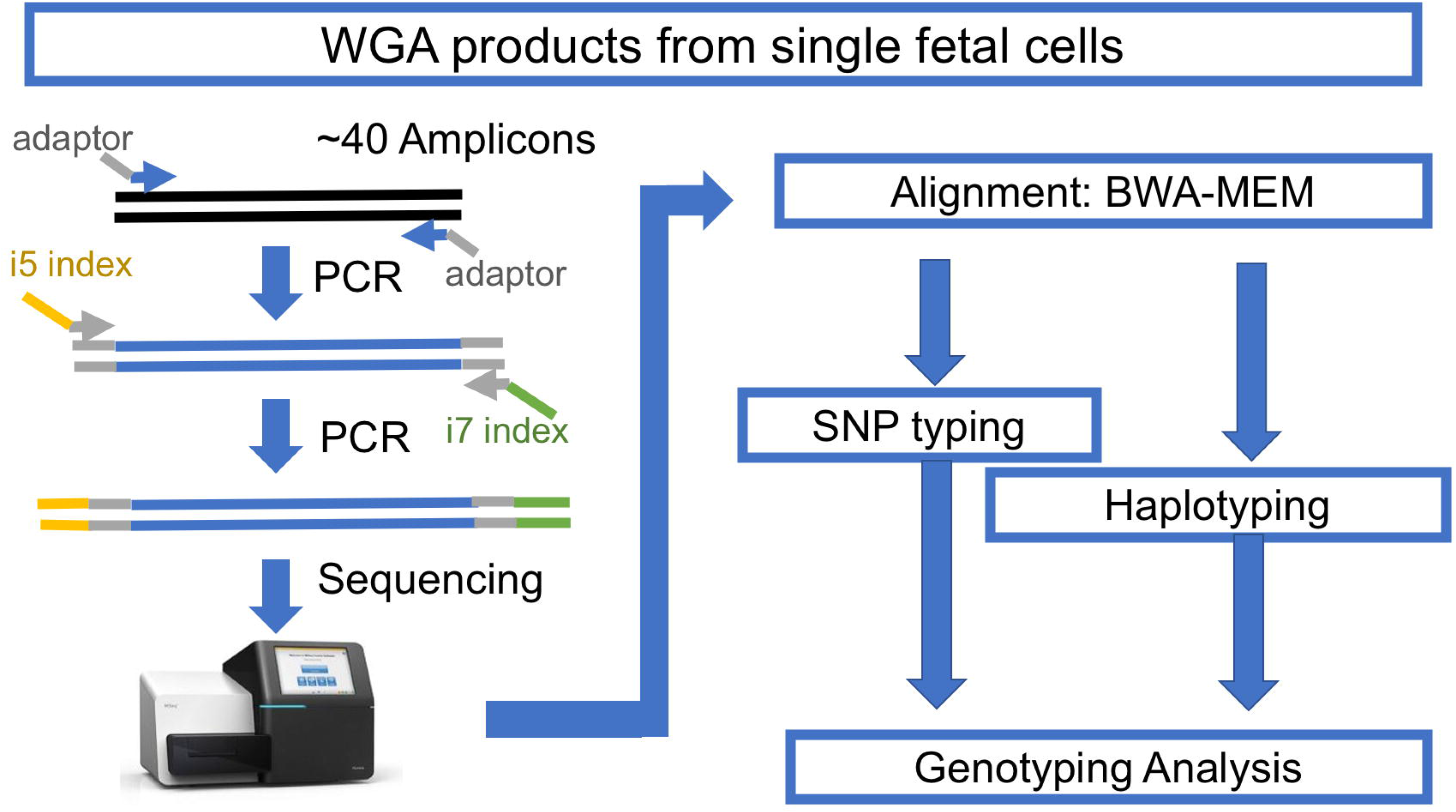
The workflow for amplicon-based genotyping.

### Amplicon design

Three groups of amplicons were used to carry out amplicon-seq. The first group consisted of amplicons with multiple common SNPs (>5% prevalence) as suggested in Debeljak et al. [8], including regions in HLA-A, HLA-B, chromosome 7q11 and chromosome 11q22, which have good sequencing coverage in single-cell WGA; all of these amplicons have at least eight common SNPs. The information of these common SNPs can be used for effective haplotyping and identifying the origin of isolated cells. The second group, which consisted of 37 amplicons, contained common SNPs selected from the Human Identification panel (ABI), which covers most chromosomes, including the Y chromosome. They were selected according to sequencing coverage in the single-cell WGA product, which has a high tendency for dropout. The third group consisted of amplicons designed to detect single nucleotide variants associated with certain inherited disease genes of interest, including hemoglobin subunit beta (*HBB*), hexosaminidase A (*HEXA*), and cystic fibrosis transmembrane conductance regulator (*CFTR*) (Table 1). For studies of trophoblasts from specific at-risk cases, we also prepared amplicons for DHCR7 and *RASPN* (Table 1). The primers of these amplicons were prepared with adaptors compatible for Illumina True-seq HT i5 and i7 adaptors.

**Table 1.** List of SNPs.

| Amplicon_ID | Chr | Amplicon_Start | Amplicon_Stop | Size | Annotation |
| --- | --- | --- | --- | --- | --- |
| rs1490413 | chr1 | 4367241 | 4367415 | 175 |  |
| rs4847034 | chr1 | 105717572 | 105717689 | 118 |  |
| **rs3780962** | chr10 | 17193291 | 17193386 | 96 |  |
| **rs964681** | chr10 | 132698373 | 132698467 | 95 |  |
| **rs1498553** | chr11 | 5708942 | 5709103 | 162 |  |
| **rs901398** | chr11 | 11096160 | 11096278 | 119 |  |
| **rs2269355** | chr12 | 6945833 | 6946005 | 173 |  |
| **rs4530059** | chr14 | 104769098 | 104769197 | 100 |  |
| **rs2016276** | chr15 | 24571747 | 24571845 | 99 |  |
| **rs2342747** | chr16 | 5868655 | 5868769 | 115 |  |
| **rs2292972** | chr17 | 80765753 | 80765827 | 75 |  |
| **rs938283** | chr17 | 77468418 | 77468592 | 175 |  |
| **rs9905977** | chr17 | 2919336 | 2919452 | 117 |  |
| **rs1024116** | chr18 | 75432299 | 75432467 | 169 |  |
| **rs576261** | chr19 | 39559774 | 39559848 | 75 |  |
| **rs12997453** | chr2 | 182413125 | 182413299 | 175 |  |
| **rs1005533** | chr20 | 39487029 | 39487201 | 173 |  |
| **rs445251** | chr20 | 15124851 | 15125009 | 159 |  |
| **rs221956** | chr21 | 43606946 | 43607048 | 103 |  |
| **rs2830795** | chr21 | 28608067 | 28608212 | 146 |  |
| **rs733164** | chr22 | 27816739 | 27816833 | 95 |  |
| **rs1355366** | chr3 | 190806053 | 190806219 | 167 |  |
| **rs4364205** | chr3 | 32417580 | 32417720 | 141 |  |
| **rs6444724** | chr3 | 193207331 | 193207425 | 95 |  |
| **rs1979255** | chr4 | 190318032 | 190318131 | 100 |  |
| **rs159606** | chr5 | 17374818 | 17374977 | 160 |  |
| **rs338882** | chr5 | 178690682 | 178690774 | 93 |  |
| **rs7704770** | chr5 | 159487871 | 159488033 | 163 |  |
| **rs13218440** | chr6 | 12059906 | 12060004 | 99 |  |
| **rs6955448** | chr7 | 4310289 | 4310423 | 135 |  |
| **rs1360288** | chr9 | 128967996 | 128968115 | 120 |  |
| **rs1463729** | chr9 | 126881396 | 126881493 | 98 |  |
| **rs7041158** | chr9 | 27985851 | 27986020 | 170 |  |
| **P256** | chrY | 8685171 | 8685289 | 119 |  |
| **rs17250845** | chrY | 8418867 | 8418960 | 94 |  |
| **rs35284970** | chrY | 2734829 | 2734921 | 93 |  |
| **rs3911** | chrY | 21733328 | 21733502 | 175 |  |
| **HLA-A** | chr6 | 2911156 | 2911140 | 245 | Haplotype |
| **HLA-B** | chr6 | 31319491 | 31319646 | 156 | Haplotype |
| **Chr7q11** | chr7 | 64895160 | 64895374 | 215 | Haplotype |
| **Chr11q22** | chr11 | 99491336 | 99491527 | 192 | Haplotype |
| **rs334&rs33930165** | chr11 | 5226980 | 5227053 | 74 | HBB |
| **rs11393960** | chr7 | 117559481 | 117559645 | 165 | CFTR |
| **rs75527207&rs74597325** | chr7 | 117587771 | 117587870 | 100 | CFTR |
| **rs121907954** | chr15 | 72350490 | 72350617 | 128 | HEXA |
| **rs387906309** | chr15 | 72346528 | 72346691 | 164 | HEXA |
| **rs147324677** | chr15 | 72346182 | 72346293 | 112 | HEXA |
| **rs138659167** | Chr11 | 71146795 | 71146921 | 127 | DHCR7 |
| rs104894299&rs761584017 | Chr11 | 47469333 | 47469720 | 388 | RASPN |

### Library construction for Illumina

WGA was performed using the PicoPLEX kits (Rubicon/Takara) or, in some cases, the Ampli1 WGA kit (Silicon Biosystems). according to the manufacturer’s protocol. Briefly, 100ng of WGA DNA product was used for Amplicon-seq library construction, with two-step PCR. The first step included 20-25 cycles of PCR with NEB Q5HS to amplify amplicons of interest and add designated adaptors. The second step used a previous adaptor sequence as primer binding sites to add Illumina i5 and i7 dual-index adaptors with 10-15 cycles of PCR. The 2-step PCR products were purified with standard AMPure protocol (Beckman) for the 100-300 bp product. PCR products were visualized by gel electrophoresis followed by the Bioanalyzer (Agilent) to check the quality and then quantified by using a KAPA Library Quantification Kit (Kapa Biosystems).

### Sequencing with Miseq

The barcoded DNA library was diluted to 2 nM and pooled together for denaturing with the Illumina protocol. An 8-10 pM diluted denature library was mixed with 5-10% PhiX control. The mixed library was loaded onto the Illumina Miseq with 150-cycle v3 kits (Illumina) and sequenced with 2×76 reads and dual index.

### Demultiplexing, alignment and variant calling

Sequencing results were demultiplexed by Illumina BaseSpace. The reads were mapped and processed with a shell script (S3 File). In summary, the fastq.gz raw files were aligned with customized reference files (S1 File) by BWA-MEM[9]. Samtools [10, 11] and Bam-readcount follow with a customized R script (S4 File) were used for calling variants within selected intervals (S2 File). The cutoff for calling a variant is at least ten reads of the less frequent allele and 5% of all reads.

Variants from each sample were summarized and compared with their paired control with an R script (S6 File). Typically, a cell with more than 2 SNPs and more than 10% of comparable SNPs different from its maternal gDNA control is considered as a likely fetal cell. Otherwise, it will be classified as an uninformative cell.

### Haplotype calling

Using the Illumina 2×76 read length necessitated performing stitch overlap for read 1 and read 2 with PEAR (Paired-end read merger) [12] for amplicons shorter than 150 bp. For amplicons larger than 150 bp, we stitched non-overlapping reads with a 15-N padding sequence. Those stitched reads were mapped with BWA-MEM with modified parameters that allow a bigger unmatched gap inside a mapped read. The mapped results were processed with an R script (S5 File) used in the main shell script to extract SNPs and reconstitute a new sequence with CIGAR information. The new concise sequences were tabulated and grouped with sequence similarity according to their Levenshtein distance. We assigned a haplotype to each major group of a concise sequence. Pair-wise haplotype comparison between the maternal gDNA sample and the putative fetal cells were performed with another R script (S7 File). All scripts are hosted and maintained on https://github.com/xmzhuo/NIPT_genotyping.

## Results

### Target region coverage

We compared the coverage of our amplicons for SNPs with gDNA and NIPT cell WGA products (Fig 2). For gDNA, most of the samples have very high coverage, which reflects the distribution of samples with many scorable SNPs. The WGA product of maternal white blood cells (WBCs) shows less scorable SNPs than gDNA, which would be the result of starting with a single diploid genome target in combination with fixation, staining, and amplification during WGA.

**Fig 2.**
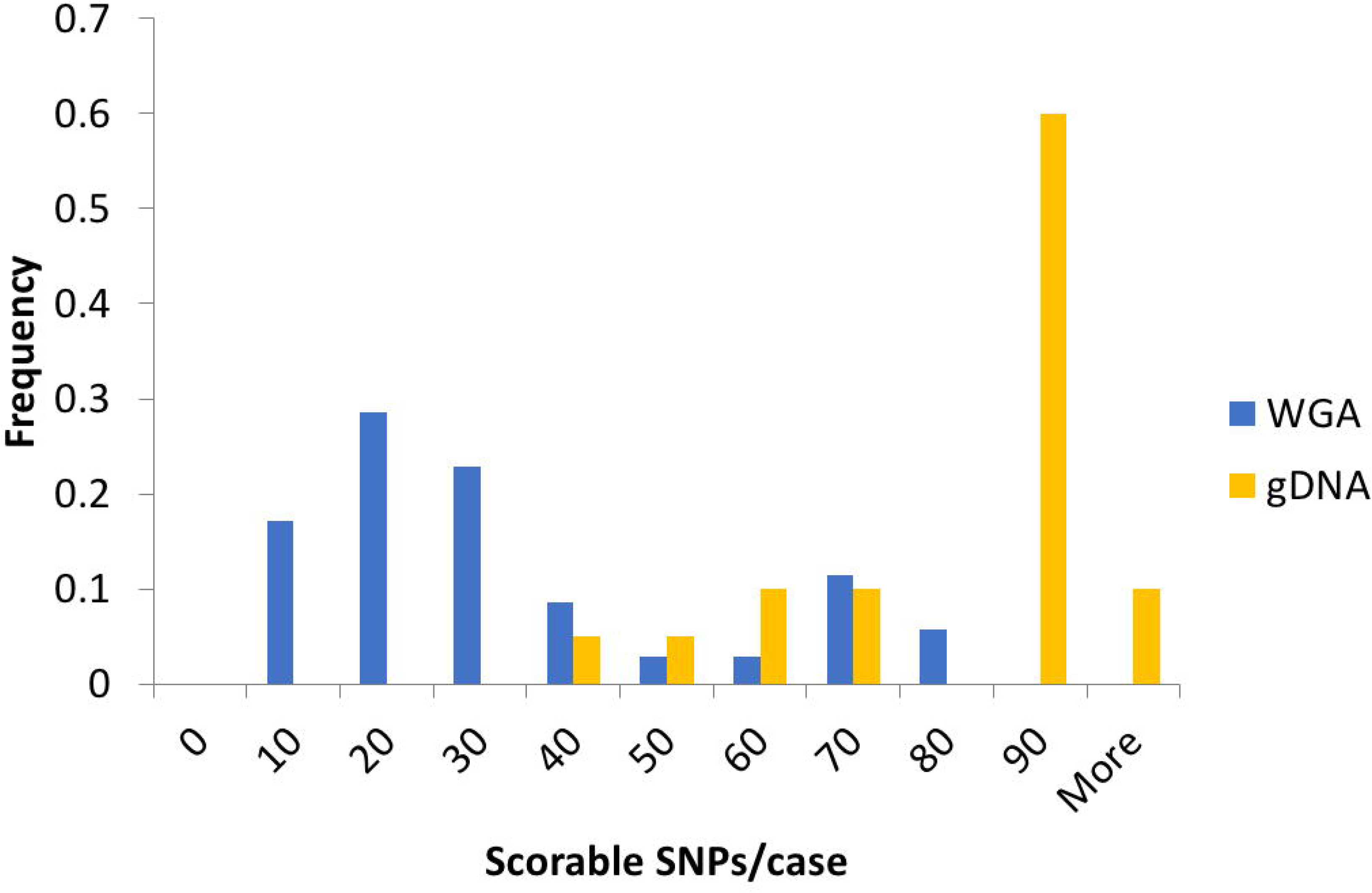
Comparison of the coverage of several genomic DNAs and NIPT single cells. The count of samples with various scorable SNPs was normalized to the total number of samples of each group. (gDNA n=20, WGA n=35). The scorable SNP cutoff is at least 5% minor allele frequency (MAF) and ten reads. Data include NIPT case numbers 946, 977, 982, 983, 984, 988, 989, 990, 991, 992, 993, 996, and 998 (clinical data in S8_File table).

### SNP typing of NIPT WGA products

A typical process of SNP typing after BWA-MEM aligner mapping includes variant calling with Samtools and retrieving the read depth with bam-readcount (Fig 3). This step will produce a table containing variant call information of allele fraction and read depth of all SNPs of interest. Information on indels was masked to avoid confusion in later steps. Then, we performed the pair-wise comparison of WGA products with the maternal gDNA. The script will calculate how many SNPs are different between two DNA samples. In an example case with two cells (Fig 3), three SNPs show the difference between the cell (middle panel) and maternal gDNA, which suggests that it is likely to be a fetal cell. The other cell shows identical calls with maternal gDNA, which suggests that it may not be a fetal cell.

**Fig 3.**
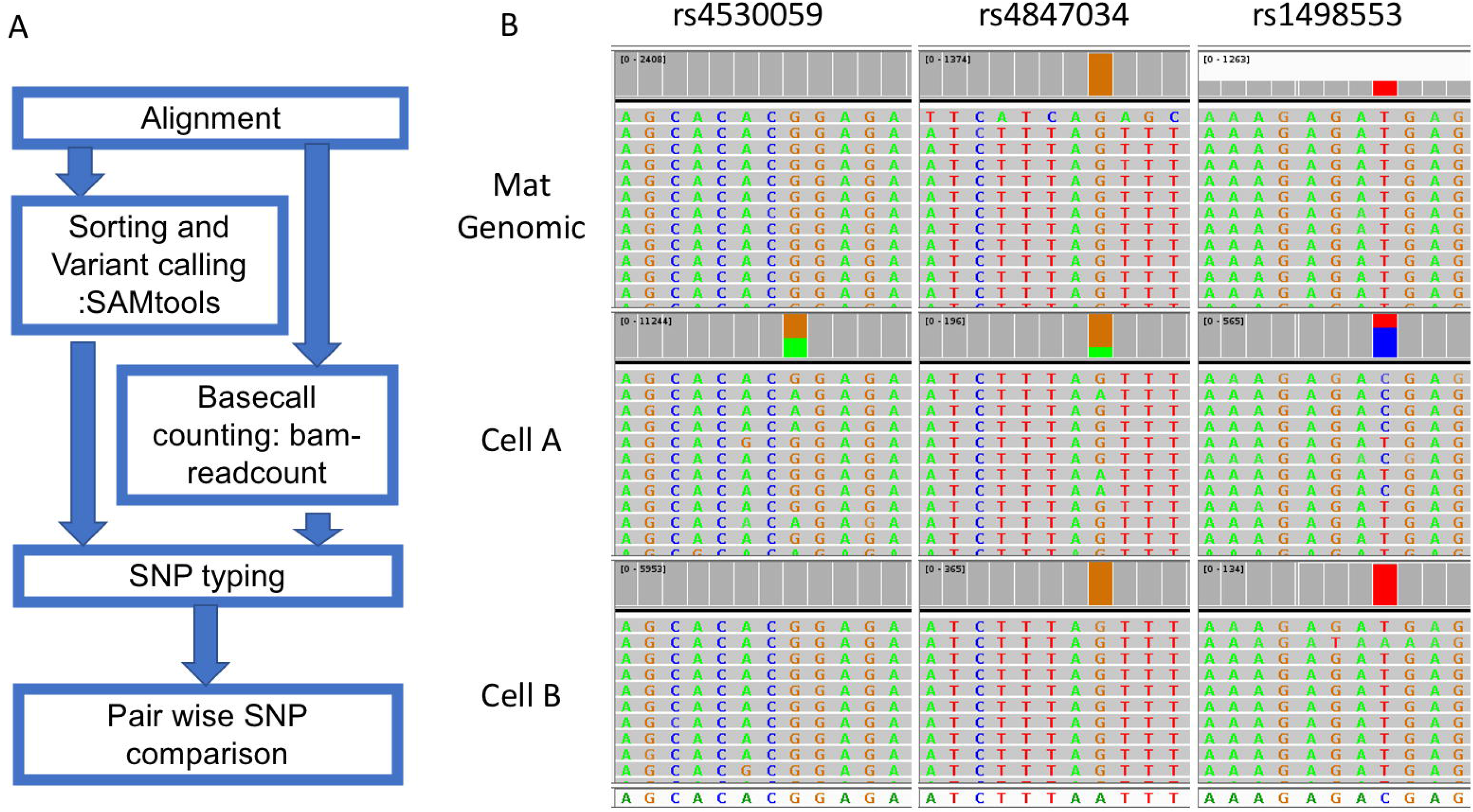
The workflow and an example of SNP typing. Reads for the mutant allele are colored green while reads for the normal allele are colored grey and summed in red. The mother is homozygous for all three SNPs, while cell A is heterozygous for all three SNPs in each case having an allele that the mother does not have. Cell B is homozygous for all SNPs indistinguishable from the mother and is interpreted as maternal or noninformative (fetal with allele drop out for all three SNPs). Data from NIPT case number 1000 (clinical data in S8_File table).

We have compared DNA from 156 blood samples, which include gDNA and DNA from WBCs and fetal cells (confirmed by orthogonal methods, such as gender PCR or low coverage WGS), to study the sensitivity and specificity of SNP typing in identifying alleles present in the cell that were not present in the mother and thus being real fetal cells. We used the WBC WGA product from the matched maternal blood as a true negative (Table 2). Typically, a SNP difference of more than 6% can largely avoid a false call in our study without missing real fetal cells. In our application, we used a SNP difference of at least two SNPs and 10% of scorable SNPs for our calling. To date, we found that 68.9% genotyped putative cells from 156 cases are informative (357/518).

**Table 2.** True positive and False positive value with various differential SNP percentage.

|  | Maternal cell | female gDNA | Fetal w/o sex | Fetal cell | WBC from<br>different<br>origin |
| --- | --- | --- | --- | --- | --- |

| diff SNP % | FPV | TPV | TPV | TPV | TPV |
| --- | --- | --- | --- | --- | --- |
| 0.01 | 0.11428571 | 0.99358974 | 1 | 1 | 0.72435897 |
| 0.02 | 0.11428571 | 0.99358974 | 1 | 1 | 0.72435897 |
| 0.03 | 0.11428571 | 0.98076923 | 1 | 1 | 0.72435897 |
| 0.04 | 0.11428571 | 0.94230769 | 1 | 1 | 0.71153846 |
| 0.05 | 0.11428571 | 0.91025641 | 0.85714286 | 1 | 0.68589744 |
| 0.06 | 0.08571429 | 0.86538462 | 0.85714286 | 1 | 0.66025641 |
| 0.07 | 0.08571429 | 0.82692308 | 0.85714286 | 1 | 0.63461539 |
| 0.08 | 0.08571429 | 0.80128205 | 0.71428571 | 1 | 0.62820513 |
| 0.09 | 0.08571429 | 0.75641026 | 0.71428571 | 0.85714286 | 0.60897436 |
| 0.1 | 0.08571429 | 0.73717949 | 0.57142857 | 0.71428571 | 0.58974359 |
| 0.11 | 0.08571429 | 0.70512821 | 0.57142857 | 0.71428571 | 0.53205128 |
| 0.12 | 0.05714286 | 0.64102564 | 0.42857143 | 0.71428571 | 0.51923077 |
| 0.13 | 0.02857143 | 0.62179487 | 0.42857143 | 0.71428571 | 0.5 |
| 0.14 | 0 | 0.55769231 | 0.28571429 | 0.71428571 | 0.48076923 |
| 0.15 | 0 | 0.53205128 | 0.28571429 | 0.57142857 | 0.41025641 |
| 0.16 | 0 | 0.47435897 | 0.14285714 | 0.42857143 | 0.37820513 |
| 0.17 | 0 | 0.41025641 | 0.14285714 | 0.28571429 | 0.36538462 |
| 0.18 | 0 | 0.39102564 | 0.14285714 | 0.28571429 | 0.33974359 |
| 0.19 | 0 | 0.35897436 | 0.14285714 | 0.14285714 | 0.28205128 |
| 0.2 | 0 | 0.32692308 | 0.14285714 | 0.14285714 | 0.23717949 |

In NIPT #1000 (Fig 4), we isolated eight putative fetal cells from the mother’s blood for WGA. Cells G78, G212, G 227, and G232 all have at least four SNPs which differed from the maternal gDNA; thus, they were confirmed to be fetal in origin. The remaining four cells (G79, G113 and G320) had 0-2 SNPs which differed from the maternal gDNA, and these were considered uninformative and possibly white blood cells accidently isolated from the mother’s blood. Cell G106 has only 2 SNP difference in less than 20 informative SNPs, which was considered as a low-quality sample and uninformative.

**Fig 4.**
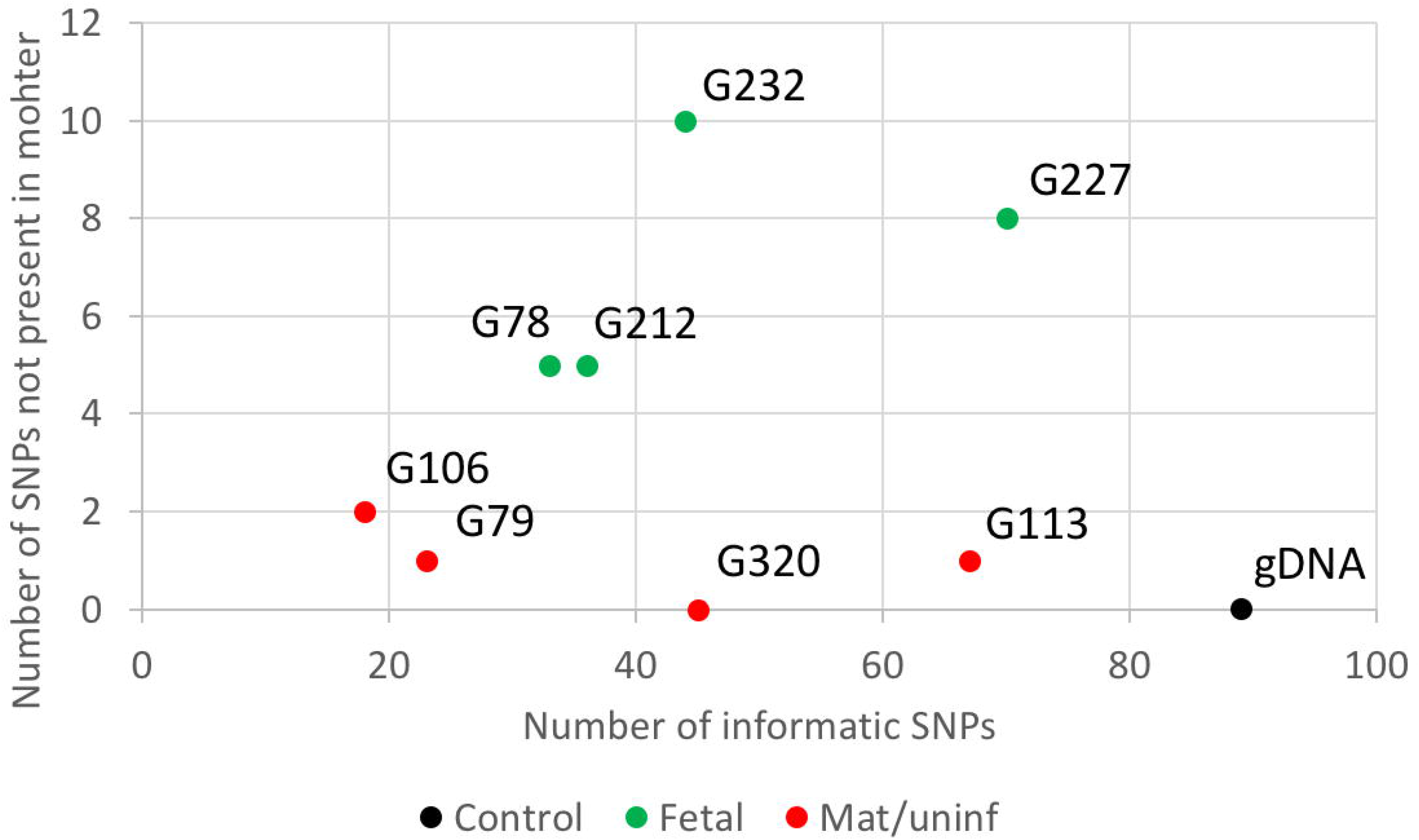
Example of SNP typing result of one NIPT case. The X-axis indicates the number of informative SNPs per sample. The Y-axis is the number of SNPs with non-maternal alleles in a cell. As expected, the maternal gDNA samples gave no alleles not present in the mother. Data from NIPT case number 1000.

### Haplotyping of NIPT WGA products

The haplotyping of NIPT WGA products potentially has higher power than SNP typing for identifying fetal cells. Since the WGA product typically went through the 14-16 cycles of amplification from trace amounts of input DNA, there is a small chance of introducing new mutations, which would affect the precision of SNP typing at low read-depth. For example, some of the cells from case #1000 (Fig 4) had one low-depth SNP difference from the maternal gDNA. To address this issue, we developed a haplotyping approach for multiple highly polymorphic regions, which contain multiple very common SNPs within the 200 bp amplicon. Thus, we can decrease the impact of random mutations introduced during the WGA process on the final interpretation (i.e., one nucleotide change is less likely to change the classification of a major haplotype group, which is comparable to HLA typing approach). Mutations introducing a random change are not rare, but mutations switching from one allele at a SNP to the other polymorphic allele are much rarer. In addition, the haplotypes of an amplicon can be treated as a permutation of a given number of SNPs, which theoretically generates much more haplotypes than SNP types and has higher power at differentiating two cells. Third, we can estimate the point at which a sequence artifact arose based on the fraction of each minor haplotype group in the total reads for a given amplicon. For example, a high fraction indicates a variant preexisted in the cell, a medium fraction indicates the variant arose during the WGA step, while a low fraction is consistent with an artifact from the final step of amplicon-seq.

We performed haplotyping with the following steps (Fig 5). After regular alignment with BWA-MEM with the default setting, we joined the Read 1 and Read 2 with PEAR [12]. The overlapping Read 1 and Read 2 were merged. If the amplicon is longer than two reads joined together, we merged the two reads and padded the gap with a tandem repeat of N. The merged reads were remapped with BWA-MEM again with a lenient setting to tolerate a larger gap. The remapped reads were processed with an R script to extract the selected SNPs, and each read was reconstituted with the concise sequence while preserving the read ID. The concise reads were then tallied and ranked according to frequency (typically, only the top 10 were kept, which usually consist of more than 99.99% of all types of reads). The Levenshtein distances were calculated for these reads, which typically ended up with only one or two major groups to represent the haplotype of this amplicon. To compare the haplotypes of more than two samples, all the top reads of each sample were pooled together, and their distances were calculated, which will determine if these samples share the same read group (haplotype). For example, the maternal gDNA carried a mocked haplotype 1 (TA) and haplotype 2 (GC). A positive fetal cell should be identified to carry at least one new haplotype 3 (GA), which would be TA/GA or GA/GA (result from dropout of haplotype 1 or 2) (Fig 5).

**Fig 5.**
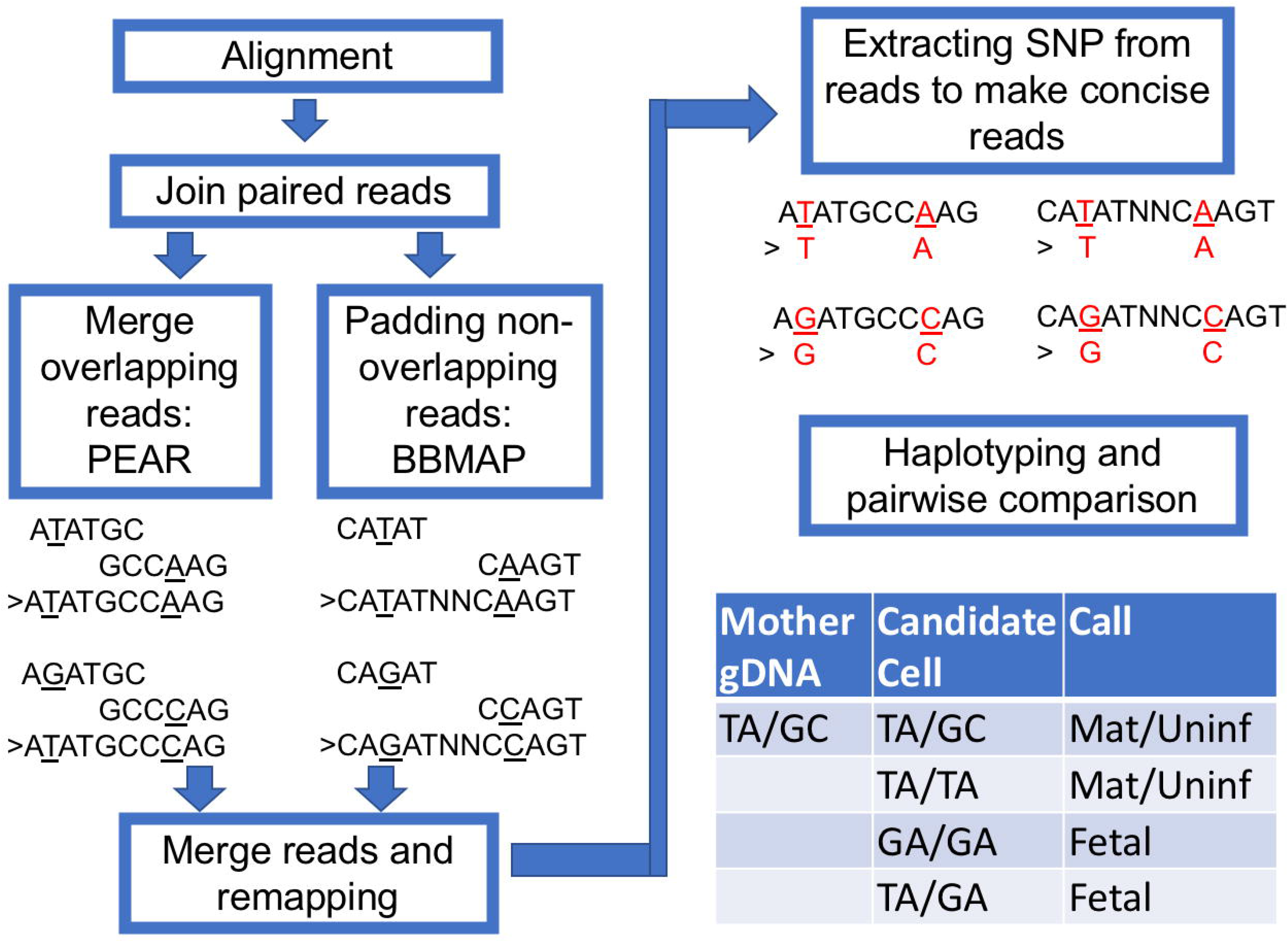
The workflow for haplotyping. The paired aligned reads were jointed with PEAR and padded with BBMap short read aligner when a gap exists. The new joint reads were remapped with BWA-MEM with a modified configuration. The highly polymorphic sites were extracted for constructing concise haplotypes. To demonstrate the workflow, we present example short reads with TA and GC haplotypes.

The haplotyping approach can effectively differentiate a candidate cell from maternal gDNA. In the case shown in Fig 6, we have the gDNA from both parents. As described previously, we extracted all 28 SNP sites in the HLA-A amplicon and reconstructed a concise 28 nt sequence for each read. In this case, the top four most frequent read types of potential fetal cells can be grouped into two major groups, with a Levenshtein distance of more than 2 (S1 Fig). The intra-group difference has a distance of less than 0.5, which suggests a difference of only one nucleotide. The difference likely results from artifacts introduced during extensive amplification (WGA then PCR). The same condition was observed in maternal gDNA and paternal gDNA as well. We observed the inheritance pattern of haplotypes when all read types from maternal, paternal, and fetal DNA were plotted together. One fetal haplotype matched with the mother and the other matched with the father. From these haplotype groupings, we concluded that this is a true fetal cell.

**Fig 6.**
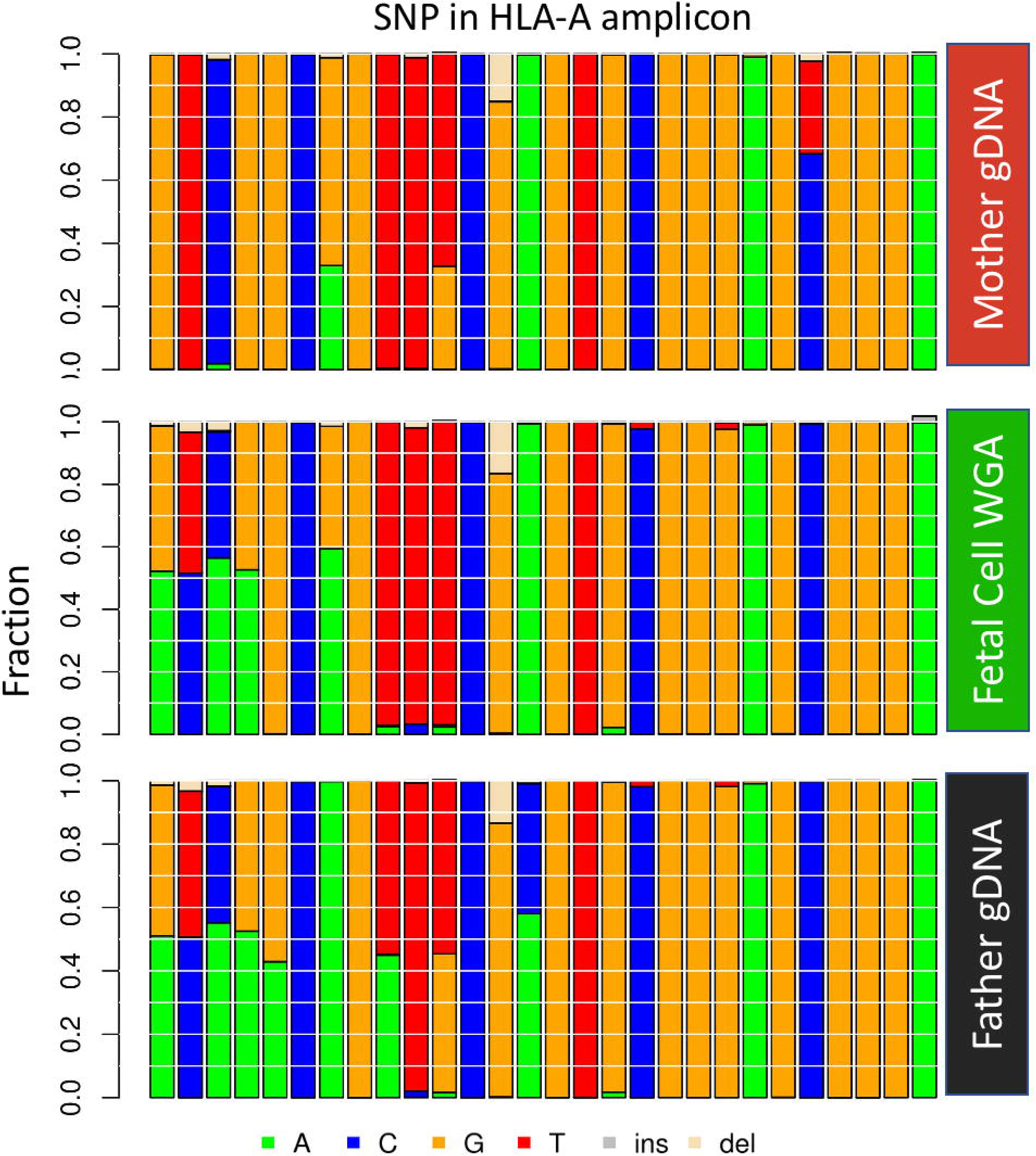
Haplotype analysis of a fetal cell compared to both parents. Each vertical column represents a particular SNP in the haplotype. The left-most four SNPs are informative as the fetal cell has an allele that the mother does not have and indicate that the putative fetal cell is indeed fetal. The 24 SNPs to the right are not informative for the fetal cell as they do not have an allele that the mother does not have. Data from NIPT case number 1000.

We tested the performance of haplotyping in four amplicons with matched gDNA, WBC, and fetal cells (S2 Fig). For gDNA, all four amplicons performed nicely to distinguish one from the other. For WBC and fetal cells, the performance was not as good, largely owing to the dropout events. However, when four amplicons were combined, we can still distinguish about 50% of all the cells. With combined power of both SNP typing and haplotyping, we can increase the solving rate of differentiating a WBC from around 60% to more than 70% (S3 Fig).

### Genotyping for monogenic disease mutations

We also wished to use this method to genotype for monogenic disease mutations. We first evaluated the ability to detect disease variants in single cultured lymphoblasts of known genotypes. Our cell based NIPT provides pure fetal DNA, which allows us to look at monogenic disease mutations. Here, we developed amplicons that contain HEXA c.805C>T, HBB c.19T>A, HBB c.20C>T, and CFTR c.1521_1523delTCT. Corresponding cells carrying certain mutations were obtained from the Coriell Institute and single lymphoblasts were picked from tissue culture and processed for WGA. The cells were isolated and genotyped by methods described herein. We successfully detected the known variants in the WGA products of cells carrying these mutations (Fig 7). The allele dropout rate was 15% and 8% for unfixed and fixed lymphobasts, respectively.

**Fig 7.**
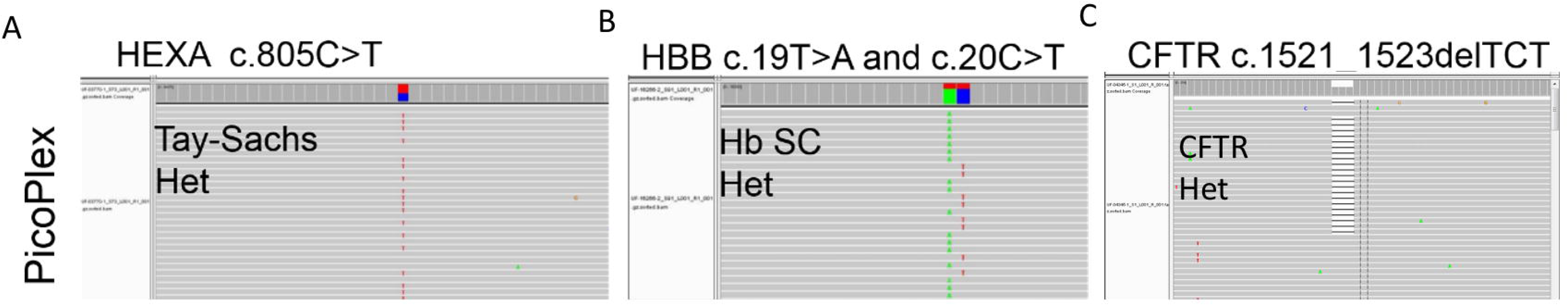
Testing for point mutations in single lymphoblasts. From left to right, Tay-Sachs, HEXA c.805C>T het; HBB c.19T>A and c.20C>T compound het; CFTR c.1521_1523delTCT het. The reads are shown in the IGV browser.

In all of our cell-based NIPT samples studies, there were three families with known monogenic mutations. One was a family where the mother was affected with sickle cell anemia, and the fetus was expected to be heterozygous based on parental information. No amniocentesis or CVS was performed. In a second family, both parents were carriers with the same known pathogenic mutation in *DHCR7*. By amniocentesis, the fetus was heterozygous for the mutation carried by both parents. A third family had a previous affected child with congenital myasthenic syndrome type 11caused by biallelic, compound heterozygous mutations in *RAPSN*. By CVS, the fetus carried the paternal but not the maternal mutation. For the sickle cell family, ten fetal cells were recovered and five were genotyped. Two cells were heterozygous for the mutation (Fig. 8), while one cell had dropout for the normal allele and one cell had dropout for the mutant allele. For the DHCR7 family, seven fetal cells were recovered and four were genotyped. One cell was heterozygous for the mutation (Fig. 8), while two cells had dropout for the normal allele and one cell had dropout for the mutant allele. For the RAPSN family, four fetal cells were recovered and four were genotyped. In Fig 8C, the mutation in each parent is shown. The G540 cell in Fig 8C, shows absence of the maternal mutation but presence of the paternal mutation. All four cells showed absence of the maternal mutation. For the paternal mutation, two cells were heterozygous while one cell had dropout for the normal allele and one cell had dropout for the paternal mutant allele. There is a small probability that the fetus carries the maternal mutation, but there was dropout in all four cells; more likely the fetus does not carry the maternal mutation in agreement with the CVS data. The fetus does carry the paternal mutation.

**Fig 8.**
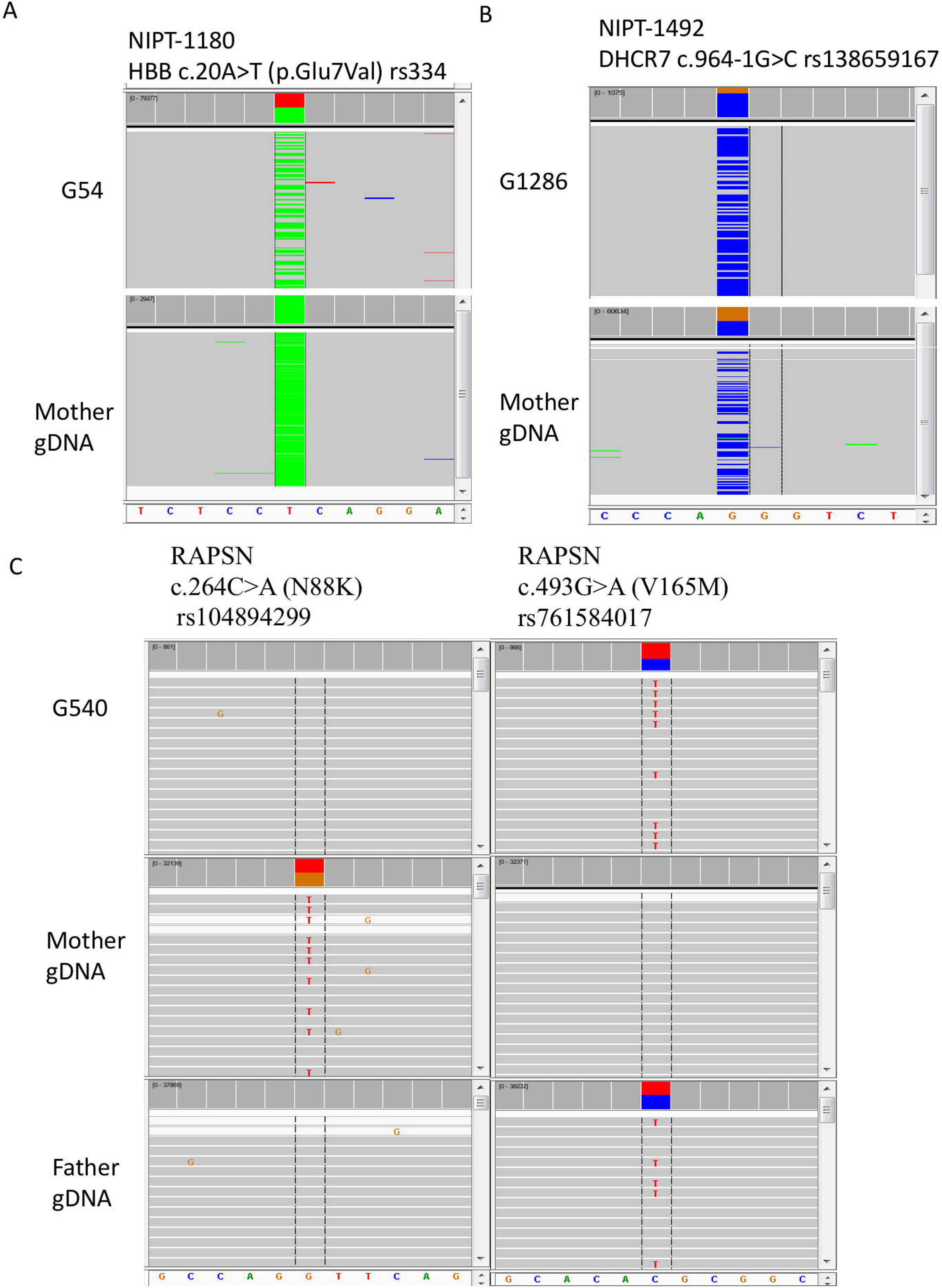
Genotyping for point mutations in trophoblasts from three cases. In panel A, the mother is affected and homozygous for the sickle mutation. Fetal trophoblast G54 is heterozygous for the mutation. Reads for the mutant allele are colored green while reads for the normal allele are colored grey and summed in red. In panel B, the mother is heterozygous for a *DHCR7* mutation that is also present in the father. Fetal trophoblast G1286 is also heterozygous for the mutation, although there is biased over-representation of the mutant allele. Reads for the mutant allele are colored blue while reads for the normal allele are colored grey and summed in brown. In panel C, the mother is heterozygous for the pathogenic N88K variant in the *RAPSN* gene, and the father is heterozygous for the V165M variant. Fetal trophoblast G540 is heterozygous for the paternal V165K variant but not for the maternal N88K variant. Allele drop out for the N88K variant cannot be ruled out, and multiple cells must be tested to gain statistical evidence that the fetus has not inherited the N88K variant. All results agreed with data from amniocentesis or CVS. Data from NIPT case numbers 1180, 1492, and 1607.

## Discussion

This single cell genotyping assay can provide an essential step for confirming the fetal origin of cells obtained from cell-based NIPT workflows. Through genotyping, we can reject cells that are of indeterminate or maternal origin and provide metrics for WGA DNA quality using multiple amplicons. Thus, researchers can focus on smaller numbers of cells, which can then be used for a more expensive downstream test or analysis, such as the low-depth NGS and microarray for CNV analysis [3, 4, 6]. We can add more amplicons to cover mutations of interest for detecting recessive or dominant inherited diseases.

Although this assay is rapid and reliable, there are still some limitations. First, the dropout rate for individual amplicons is significant, which hinders the power of cell identification and affects the detection of disease-causing mutations. Second, although the cost is relatively low and the running time is short (<24 h), it still takes extra effort to complete and could increase the turnaround time of NIPT. Third, the multiple steps of PCR after WGA are prone to introduce errors that may cause ambiguity at SNP typing, although errors switching from one SNP allele to another or from a mutant to wild-type genotype or vice versa are very rare.

There are multiple options for reducing the dropout rate for the improvement of this assay. First, we can increase the size of the panel, which allows more amplicons to compensate for the dropout. Second, we can modify the amplicons to reduce the size of the amplicons or otherwise improve the amplification. Third, improved versions of WGA are being developed which can reduce allele dropout.

To minimize the errors introduced by the multiple amplification steps, we can incorporate the genotyping amplification into WGA. We can redesign our genotyping primers to make them compatible with the linear amplification step in WGA, which will result in the enrichment of our regions of interest. The modified WGA product can be used for barcoding directly for low-depth WGS. This approach may eliminate 5-10 cycles of PCR and reduce the chance of dropout.

Use of this method for genotyping fetuses at risk for specific monogenic mutations is feasible. Although cell-free NIPT is relatively straight forward for genotyping paternal mutations, it is more complex for determining the maternal contribution to the genotype, although this can be accomplished with more complex analysis [14, 15]. Cell-free NIPT has been used to screen for *de novo* mutations in a panel of genes [13]. Confirmation for *de novo* mutations can be performed by specific reanalysis of maternal plasma if the mother is not mosaic for the mutation. Single cell analysis of circulating trophoblasts can be used to determine the genotype of fetuses at risk for monogenic disorders, although allele dropout must be ruled out by analysis of multiple cells, and a low failure of diagnosis rate is to be expected due to failure to isolate fetal cells and/or to allele drop out for cells isolated.

Our genotyping assay has the potential to be used in many clinical research applications. As described previously, it can be used to identify a fetal cell in cell-based NIPT and screen for known disease-causing point mutations. It also has the potential to distinguish between same sex dizygotic twins. Furthermore, this technique would also be adapted for other single cell applications, such as circulating tumor cell analysis. An improved version of this assay may significantly reduce the dropout rate in amplicons and increase the coverage at regions of interest.

## Data Availability

All relevant data are within the manuscript and its Supporting
Information files. Raw data is available upon request according to IRB requirement.

https://github.com/xmzhuo/NIPT_genotyping

## Acknowledgment

This work was supported by internal institutional funds at Baylor College of Medicine.

We thank the participating patients, study coordinators, and genetic counselors at Baylor College of Medicine, Texas Children’s Hospital, and Columbia University Medical Center for recruitment of samples.

## Supplement Data

**S1_File Reference sequence fasta file**

**S2_File SNVs reference BED**

**S3_File Linux shell script and parameters for alignment and analysis**

**S4_File R script for genotype calling**

**S5_File R script for haplotype calling**

**S6_File R script to compare genotype of NIPT cells with parents**

**S7_File R script to compare haplotype of NIPT cells with parents**

**S8_File Table of Clinical Sample Information**

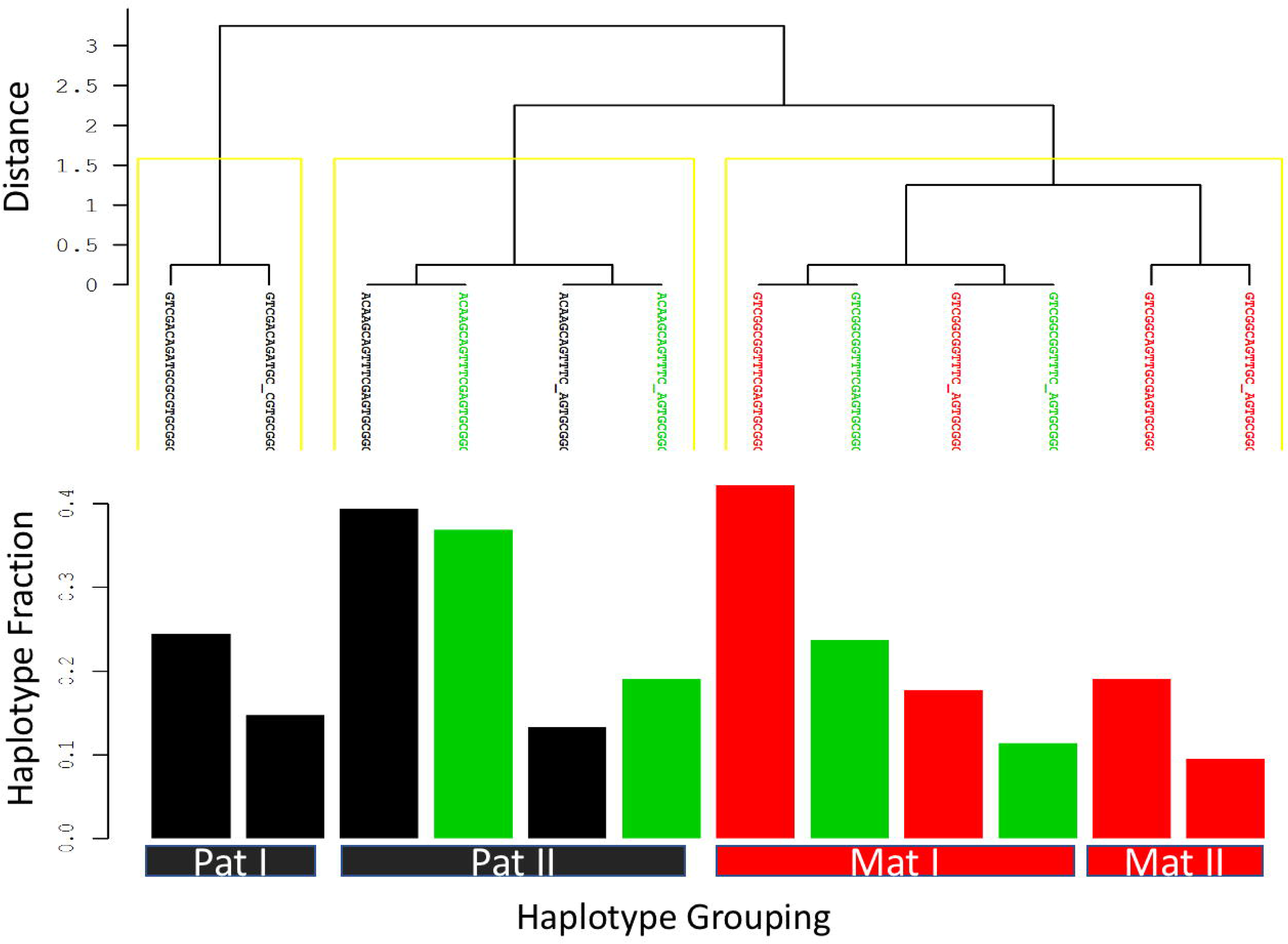

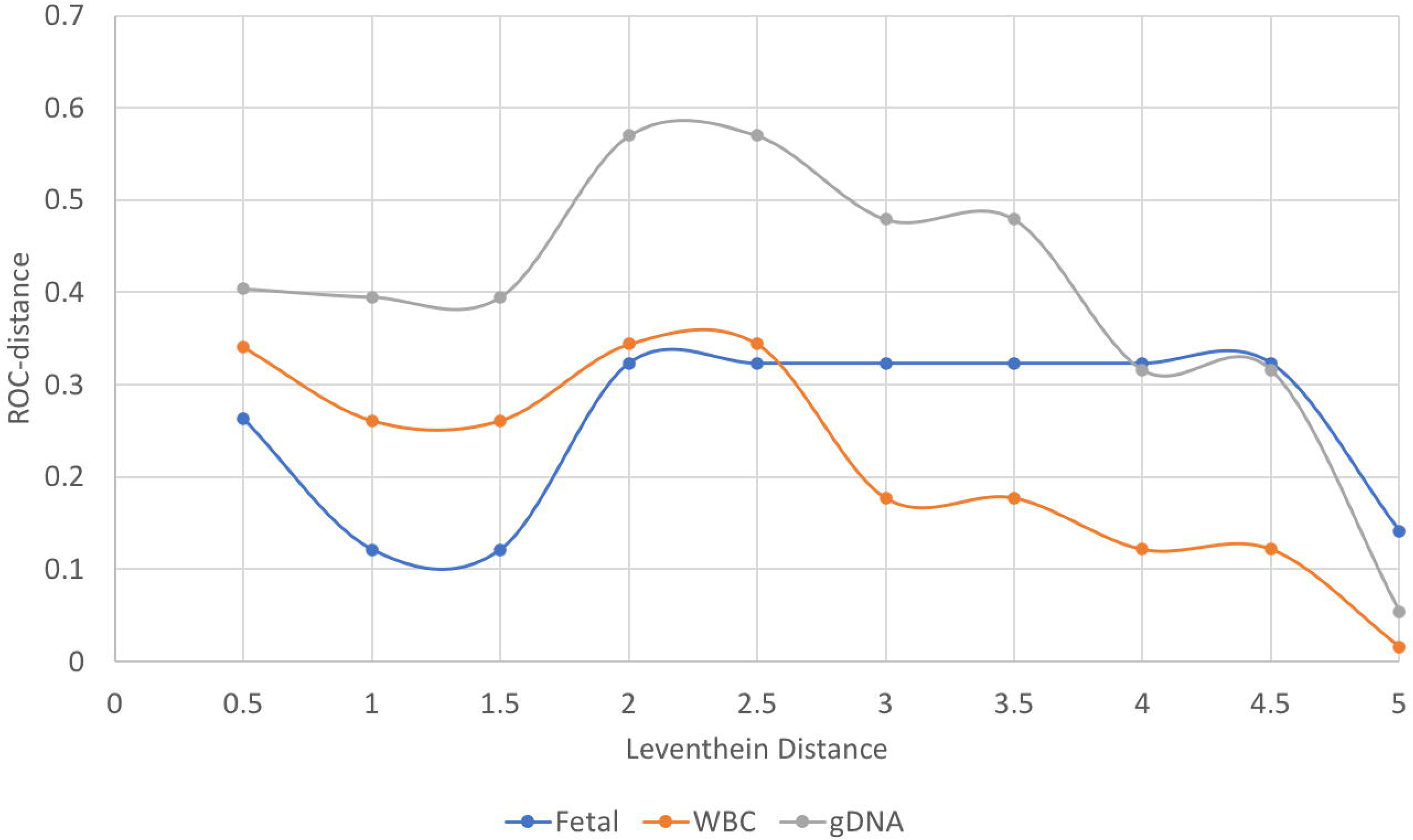

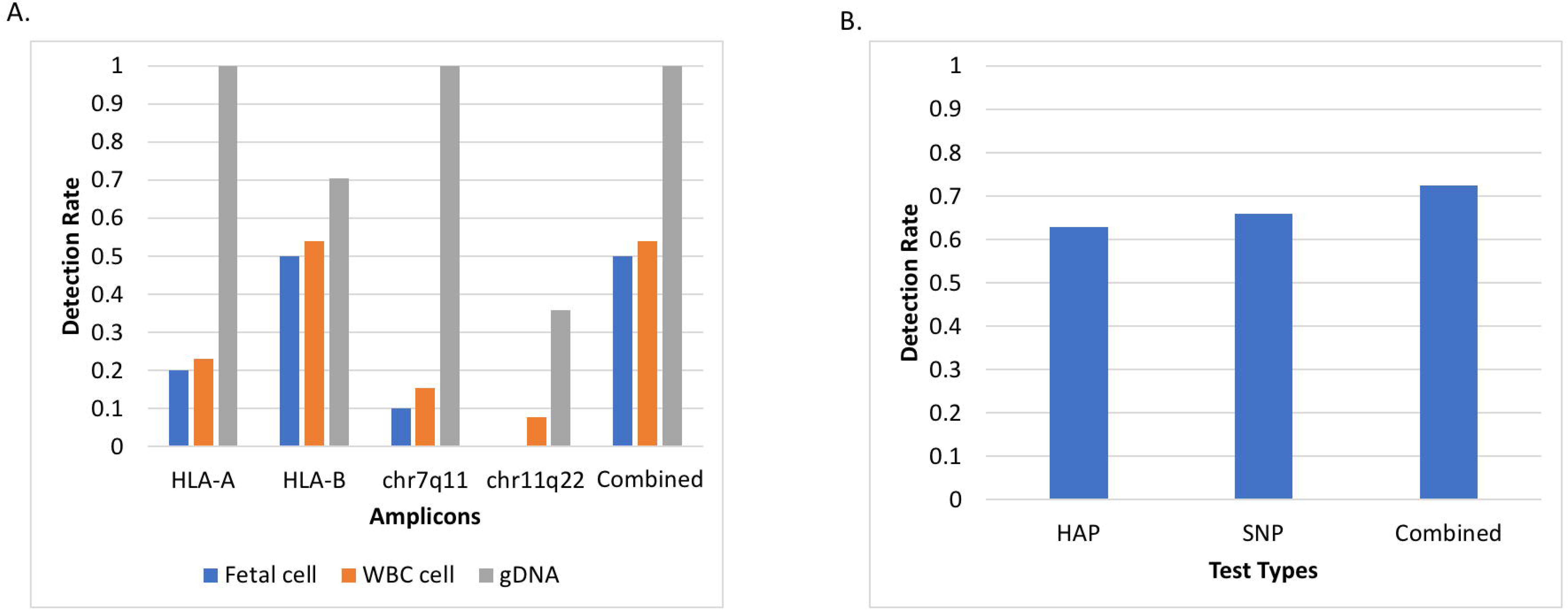

